# Predictive value of automated coronary calcium scoring in lung cancer screening with low dose computed tomography

**DOI:** 10.1101/2022.06.13.22276338

**Authors:** Federica Sabia, Maurizio Balbi, Roberta E. Ledda, Gianluca Milanese, Margherita Ruggirello, Camilla Valsecchi, Alfonso Marchianò, Nicola Sverzellati, Ugo Pastorino

## Abstract

Coronary artery calcium (CAC) is a known risk factor for cardiovascular events, but not yet routinely evaluated in Low Dose Computed Tomography (LDCT) screening. The present analysis compared the accuracy of a new automated CAC quantification versus prior manual quantification on baseline LDCT screening images as predictors of all-cause mortality at 12 years.

The study included 1129 volunteers of the Multicentric Italian Lung Detection (MILD) trial who underwent a baseline LDCT scan from September 2005 to September 2006, already analyzed in a previous paper on CAC scoring. The initial manual CAC (mCAC) had been scored by one operator using a dedicated software, while the new automated CAC (aCAC) score was measured by a fully automated artificial intelligence software. All CAC scores were stratified in four categories: 0, 0.1- 19.9, 20-399, and ≥ 400.

The study showed a high correlation between aCAC and mCAC scores, with an Intraclass Correlation Coefficient of 0.887. Of 613 negative mCAC score, 87.6% had aCAC score >0, and 14.0% >20. A CAC score >20 revealed a higher risk of 12-year all-cause mortality both with mCAC and aCAC. Focusing on the 535 individuals with false negative mCAC score, aCAC identified a subset of volunteers with a significantly poorer survival of 86% (aCAC 20-399, p=0.0007).

CAC quantification could be accurately and safely performed with a fully automated software on baseline LDCT screening images to predict all-cause mortality risk.

## Introduction

Coronary artery calcium (CAC) is an independent predictor of cardiovascular (CV) events (1). Previous studies in low dose computed tomography (LDCT) screening participants demonstrated the predictive value of CAC score for all-cause and CV mortality (2,3), with a CAC scoring accuracy of LDCT that was similar to the one achieved by electrocardiographic-gated cardiac CT (4,5).

CAC evaluation is not routinely performed in LDCT screening because manual CAC measurements is a highly time-consuming procedure, but currently available artificial intelligence (AI) software allows a fully automated quantification with high accuracy (6).

The aim of the present study was to compare the accuracy of an automated CAC quantification versus manual quantification on baseline LDCT (3) and assess their predictive value for all-cause mortality at 12 years.

## Material and Methods

The study population is represented by the subset of participants of the Multicentric Italian Lung Detection (MILD) trial volunteers (ClinicalTrials.gov Identifier: NCT02837809) who underwent a baseline LDCT scan from September 2005 to September 2006 analysed in a previous paper on manual CAC evaluation (3).

LDCTs were acquired by a 16–detector row CT scanner (Somatom Sensation 16; Siemens Medical Solutions, Forchheim, Germany): the whole chest volume was scanned during one deep inspiratory breath-hold, without the use of contrast medium and with the following scanning parameters: tube voltage, 120 kV; effective tube current, 30 mAs; individual detector collimation, 0.75 mm; gantry rotation time, 0.5 second; and pitch, 1.5. Neither electrocardiographic triggering nor dose-modulation systems were used. Images were reconstructed as follows: one-millimeter-thick sections were reconstructed with an increment of 1 mm (medium-sharp kernel, B50f), and 5-mm-thick sections were reconstructed with an increment of 5 mm (medium-smooth kernel, B30f).

The manual CAC evaluation (hereafter named as mCAC) has been described previously (3); briefly, CT images were transferred to a workstation (Leonardo; Siemens Medical Solutions) and analyzed by one operator with 5 years of experience in cardiac imaging. mCAC was performed on the 5-mm-thick images dataset using a dedicated software (CaScore; Siemens Medical Solutions) (3).

For the automated evaluation, 1-mm images were transferred to a dedicated graphic station (Alienware Area 51 R6 equipped with Dual NVIDIA GeForce RTX 2080 OC graphics) and analyzed using a fully automated AI software (AVIEW, Coreline Soft). Automated CAC (aCAC) was measured with a scoring tool based on a 3-dimensional U-net architecture.

The vital status was obtained through the Istituto Nazionale di Statistica (ISTAT, SIATEL 2.0 platform). Participants accumulated person-years of follow-up from the date of baseline until death or the date of the last follow-up as of November 2021. Both mCAC and aCAC scores were stratified in four categories: 0, 0.1-19.9, 20-399, and ≥ 400. Categorical variables were reported as numbers and percentages, whereas continuous variables as medians with interquartile ranges (IQRs); associations were evaluated by the chi-square test for categorical data and by the Mann-Whitney U test for continuous variables. Correlation between mCAC and aCAC categories was estimated by the Intraclass Correlation Coefficient (ICC) with 95% Confidence Interval (CI). Kaplan-Meier curves for 12-year all-cause survival were reported (a) in strata of mCAC score in all participants, (b) in strata of aCAC score in all participants, and (c) in strata of aCAC score (0.1-19.9 vs. 20-399) among negative mCAC score participants. Comparisons were tested by Log-Rank test for trend. Analyses were performed using the Statistical Analysis System Software (Release SAS:9.04; SAS Institute, Cary, North Carolina, USA) and R Statistical software (R Studio).

## Results

Of the initial cohort of 1159 participants, 30 (2.6%) failed the automated AI software evaluation due to LDCT features. Final study population consisted in 1129 participants: median age was 57 years, 68% were males and 65% were current smokers (Table 1).

**Table 1.** Characteristics and outcomes of study population according to Automated CAC Scores.

|  |  | Total | Automated CAC Score |  |  |  | p-value |
| --- | --- | --- | --- | --- | --- | --- | --- |
|  |  |  | 0 | 0.1-19.9 | 20-399 | 400+ |  |
|  |  | 1129 | 90 (8.0%) | 546 (48.4%) | 393 (34.8%) | 100 (8.9%) |  |
| **Age** | **Median (IQR)** | 57 (53-61) | 54.5 (51-59) | 55 (52-59) | 59 (54-63) | 61 (56-66.5) | <0.0001 |
| **Sex** | **Male** | 770 (68.2%) | 44 (5.7%) | 312 (40.5%) | 321 (41.7%) | 93 (12.1%) | <0.0001 |
|  | **Female** | 359 (31.8%) | 46 (12.8%) | 234 (65.2%) | 72 (20.1%) | 7 (2.0%) |  |
| **Smoking Status** | **Ex Smoker** | 396 (35.1%) | 36 (9.1%) | 178 (45.0%) | 139 (35.15) | 43 (10.9%) | 0.1578 |
|  | **Current Smoker** | 733 (64.9%) | 54 (7.4%) | 368 (50.2%) | 254 (34.7%) | 57 (7.8%) |  |
| **mCAC Score** | **0** | 613 (54.3%) | 76 (12.4%) | 449 (73.3%) | 86 (14.0%) | 2 (0.3%) | <0.0001 |
|  | **0.1-19.9** | 160 (14.2%) | 4 (2.5%) | 61 (38.1%) | 95 (59.4%) | 0 |  |
|  | **20-399** | 279 (24.7%) | 10 (3.6%) | 34 (12.25) | 195 (69.9%) | 40 (14.3%) |  |
|  | **400+** | 77 (6.8%) | 0 | 2 (2.6%) | 17 (22.1%) | 58 (75.3%) |  |
| **12-year all-cause mortality** |  | 100 (8.9%) | 3 (3.3%) | 28 (5.1%) | 45 (11.5%) | 24 (24.0%) | <0.0001 |

aCAC score was 0in 90 participants (8.0%), 0.1-19.9 in 546 (48.4%), 20-399 in 393 (34.8%), and ≥ 400 in 100 cases (8.9%); mCAC score was 0 in 613 participants (54.3%), 0.1-19.9 in 160 (14.2%), 20-399 in 279 (24.7%), and ≥ 400 in 77 cases (6.8%).

Subjects with aCAC score ≥20 were older (p-value <0.0001) and the frequency of aCAC ≥20 was significantly higher in males than in females (12% of males had aCAC ≥ 400 compared to 2% of females). Non-significant differences were observed for smoking status (Table 1).

The comparison of aCAC with mCAC scores, showed a high correlation (ICC = 0.887 (0.874-0.899). Of 613 negative mCAC score, 87.6% had aCAC score >0, and 14.0% >20. The 2 cases ≥ 400 proved to be failures of AI calculation.

Twelve-year all-cause mortality was 8.9% overall (100/1129), 3.3% with aCAC = 0, 5.1% = 0.1-19.9, 11.5% = 20-399, 24% = ≥ 400, 4.9% with aCAC <20, and 14.0% with aCAC > 20.

Survival curves show that participants with CAC score >20 had a higher risk of 12-year all-cause mortality, both with mCAC scoring (Figure 1A) and with aCAC scoring (Figure 1B), but the high-risk group was 44% of the cohort with aCAC vs. 32% with mCAC. Volunteers with aCAC score <20 had a much better survival than subjects with aCAC score ≥20 (95% vs. 86%, Log-rank p-value<0.0001) or ≥400 (95% vs. 76%, Log-Rank p-value<0.0001). Focused analysis of 535 individuals with negative mCAC and positive aCAC score (Figure 1C), showed that aCAC identified a subset of 86 volunteers with a significantly poorer survival of 86% (aCAC 20-399, p=0.0007).

**Figure 1.**
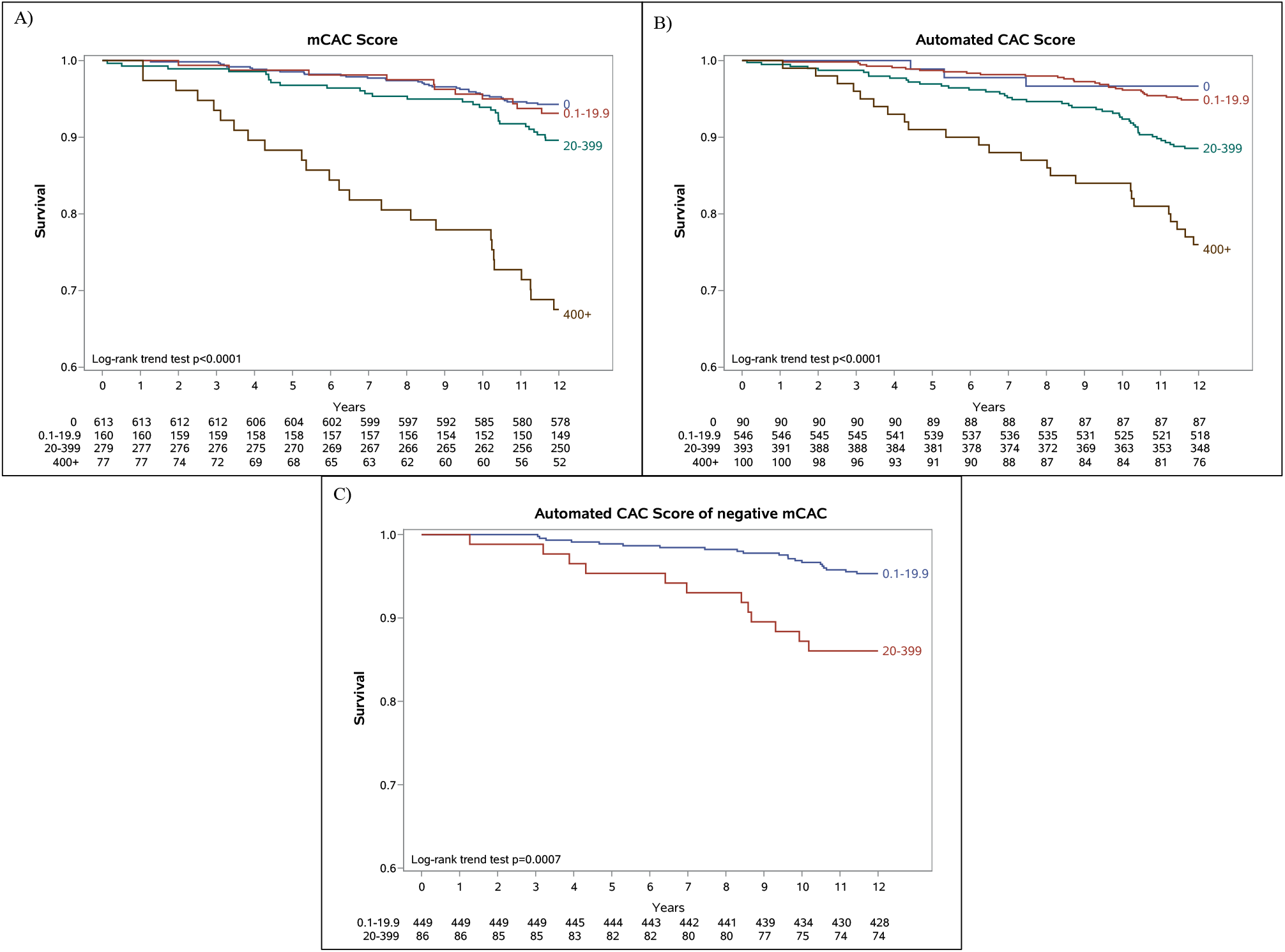
Kaplan-Meier survival curves for all-cause mortality: A) stratified by mCAC score in all 1129 participants; b) stratified by aCAC score in all 1129 participants; and C) stratified by aCAC score in participants with negative mCAC score and positive automated aCAC score.

## Discussion

Our findings showed that CAC quantification can be accurately performed with a fully automated software. The correlation between the two measurements was high (ICC of 0.887), despite the automatic software tended to overestimate CAC (12). In addition, manual assessment of CAC classified 54% of volunteers as negative, while only 8% were classified as negative by the automated score.

The automated AI-driven analysis of coronary artery calcifications of MILD volunteers confirmed our previous observation that CAC is an independent predictor for all-cause mortality (3), and that CAC > 400 is associated with a significantly worse survival at 12 years. These results are in keeping with a case-cohort study from the NELSON trial demonstrating that higher strata of CAC score had a significantly higher risk of coronary events compared with negative (10), and with a meta-analysis of 6 LCS trials showing that subjects with high CAC values had more than two-fold increased relative risk of all-cause mortality (11). The extended follow-up of MILD trial revealed a 3-fold higher mortality risk for aCAC >20 and 5-fold risk for aCAC >400 (p<0.0001 and p<0.0001 respectively). As reported in Figure 2C, aCAC stratified false negative mCAC volunteers in two categories with significantly different risk profile.

AI has been increasingly used in diagnostic radiology and different reports have compared fully or mainly automated CAC scoring vs. manual quantification, showing promising results (6, 13-15). A recent study demonstrated that a deep learning-based automated CAC assessment could be reliably used for CV risk stratification on non-gated chest CT, with both 1- and 3-mm reconstruction (16). Major advantages of an automated CAC quantification include a potential increase in measurement reproducibility, while reducing the time-consuming process of manual coronary arteries segmentation.

This study has few limitations. First, the retrospective design is prone to confounding factors, such as selection of patients. Second, images reconstruction thickness might have affected the categorical agreement. Indeed, CAC quantification on 1-mm LDCT is known to overscore calcium as compared to 5-mm LDCT, suggesting that part of the CAC overestimation from AI analysis might be due to different slice thickness rather than to a true software misclassification.

In conclusion, a fully automated quantification of the CAC score by means of an AI software could be safely performed on chest LDCTs for the purpose of mortality risk stratification within lung cancer screening activities.

## Data Availability

All data produced in the present study are available upon reasonable request to the authors

## Funding sources

The MILD trial was supported by grants from the Italian Ministry of Health (RF 2004), the Italian Association for Cancer Research (AIRC 2004 IG 1227 and AIRC 5xmille IG 12162), Fondazione Cariplo (2004-1560), and the National Cancer Institute (EDRN UO1 CA166905). The sponsors had no role in conducting and interpreting the study.

## Notes

### Competing Interest Statement

The authors have declared no competing interest.

### Clinical Trial

NCT02837809

### Clinical Protocols

https://www.clinicaltrials.gov/ct2/show/NCT02837809?term=NCT02837809&draw=2&rank=1

### Author Declarations

The Institutional Review Board and the Ethics Committee of the Fondazione IRCCS Istituto Nazionale dei Tumori of Milan approved the study.

